# Longitudinal immunophenotyping to track motor progression in Parkinson’s Associated with a TH mutation

**DOI:** 10.1101/2024.01.03.23300647

**Authors:** A. Gopinath, A. Ramirez-Zamora, S. Franks, T. Riaz, A. Smith, G. Dizon, L. Hornstein, J. Follett, C. Swartz, J. Bravo, E.L. Kugelmann, M. Farrer, M.S. Okun, H. Khoshbouei

## Abstract

Background and Objectives: PD is the second most common neurodegenerative disorder and the fastest growing. Genetic factors account for ∼15% of cases. Despite some consistency in symptoms across idiopathic and genetic PD cases, tracking progression and treatment response remains an important challenge especially in the development of new therapies. There have been many traditional approaches to tracking including DaTscan imaging, cardiac 123I-MIBG scintigraphy, MRI, CSF analysis, and following clinical symptom progression. Methods: Our previous work showed that peripheral blood mononuclear cells (PBMCs) expressing dopamine transporter (DAT) and tyrosine hydroxylase (TH) in PD patients may correlate with disease progression and with the response to treatment with levodopa. We describe a single case longitudinal follow up of a 40-45-year-old woman with PD who carried a heterozygous TH mutation. We assessed her clinical features over 18 months with DaT scans and immunophenotyping of her PBMCs. Her data were compared with idiopathic PD (n=130 subjects, both sexes) and healthy controls (n=80, age/sex matched). Results: The results revealed a rise in DAT+ immune cells which occurred coincident to documented worsening of her UPDRS-III motor scores. Unlike idiopathic PD patients, following levodopa therapy, the TH+ immune cell levels remained elevated, despite UPDRS-III score improvement. Discussion: The longitudinal immunophenotyping in this PD patient with a TH mutation suggested that DAT+ and TH+ PBMCs could be candidate biomarkers for PD progression and possibly treatment effectiveness. This study provides proof of concept to explore this approach to investigate immunophenotyping in PD progression.

## Introduction

Parkinson’s disease (PD) is the second most prevalent neurodegenerative syndrome and is the fastest growing^1^. PD is not one disease, and thus the pathophysiology across known causes is complex. Genetic factors account for ∼15% of PD cases^2,3^ with several known pathogenic gene mutations ^4,5^. Many of the symptoms and some of the progression patterns in PD are similar, whether idiopathic or genetic^6,7^. Currently, tracking the progression of PD or its response to symptomatic treatments, such as levodopa, has been attempted using imaging modalities such as DAT and MRI, as well as skin biopsy and MIBG cardiac testing^8–13,14^. While recent research has highlighted several promising disease progression markers^15–17^, their longitudinal applicability in monitoring progression for individual PD patients has not been clearly demonstrated.

We previously studied peripheral blood immunophenotyping as an approach to disease progression. We used a cross-sectional design with 130 PD patients and the data revealed that peripheral immune cells expressed both a marker for dopamine transporter (DAT+) and tyrosine hydroxylase (TH+) in idiopathic PD^17^. The levels of DAT and TH decreased following treatment with levodopa and coincident improvement in UPDRS-III (PD motor) scores^17^. Herein, we report the case of a single PD subject who carried a heterozygous TH mutation and we performed immunophenotyping to determine the consistency of the findings with idiopathic PD.

## Results & Discussion

### Patient information and Diagnostic

The patient was a 40-45 year old female who presented with a right-hand resting tremor, bradykinesia and rigidity which slowly progressed over a period of two years. She had a long history of constipation and hyposmia prior to the development of her motor symptoms. Her symptoms progressed with increased bradykinesia, shuffling gait, lower extremity pain, dystonia and impaired hand dexterity. She was initially treated with a dopamine agonist due to her age and risk of motor complications. Because of a lack of meaningful benefit and persistent nausea, carbidopa/levodopa was initiated. Her levodopa dose was titrated over time to 700 mg/day in three divided doses. She showed marked, clear and sustained response to levodopa and improvement in her motor symptoms. Her initial motor UPDRS score prior to initiation of therapy was 27, and this was reduced to a motor score of 10 at 24 months following optimization of her medical therapy. DaTscan’s were collected at baseline and at 11 months. The series of DaTscans revealed a pattern consistent with parkinsonism (Figure 1). Methods are provided in Supplementary Material: DaTscan.

**Figure 1.**
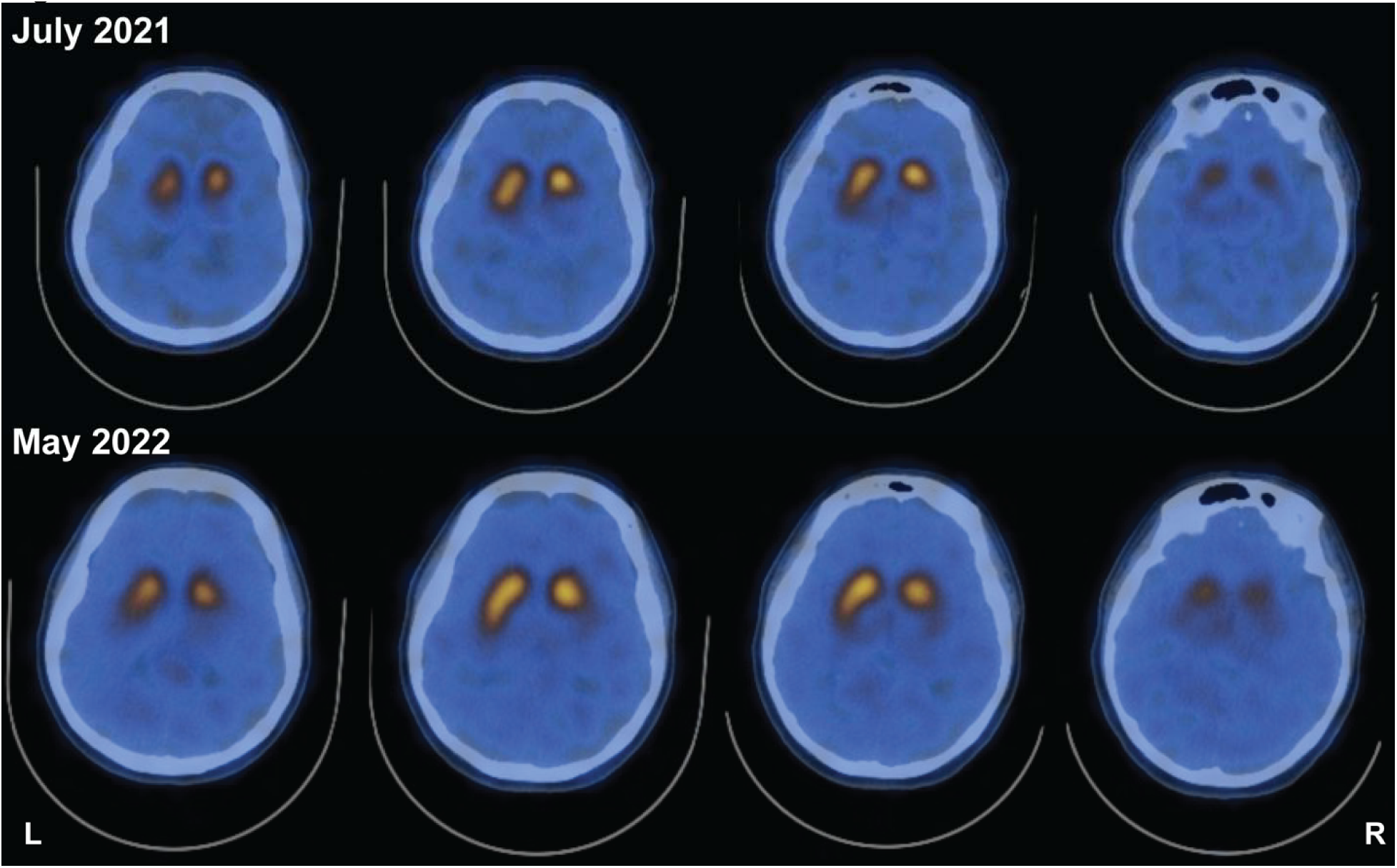
DaTscan reveals left-side asymmetry in striatal DAT binding and is not qualitatively different 11 months after initial scan. Study showing reduced striatal DAT binding both at study initiation and 11 months afterwards. Despite worsened symptoms and clinical presentation, two scans 11 months apart do not reveal changes in asymmetry.

### Sequencing

Whole genome sequencing was completed at a median 32X coverage (63% bases >30X) by Illumina 150bp paired-end reads. All known genes for parkinsonism were examined. The patient was found to be a heterozygous carrier of a pathogenic variant in *TH* (GRCh37/hg19 chr11:2,186,975-6; ENST00000381178 c.1215_1216delCCinsTT; p.Glu406*; CADD v1.6 = 49) The sequencing depth at this position was 62 reads (Ref F:R 15:20, Alt F:R 18:9). The global minor allele frequency of this mutation in control individuals was 1.59E-06 (1/627,326 alleles) albeit with a frequency of 2.8E-05 in East Asia (1/36,048 alleles; gnomAD v4.0.0). A detailed inspection of read alignment, including all single nucleotide and structural variants in the locus, did not identify any other potentially damaging variants in the gene, and this was consistent with the patient possessing at least one normal *TH* allele. TH mutations have been associated with autosomal recessive Segawa syndrome, an infantile/juvenile onset dopa-responsive dystonia-parkinsonism disorder. Heterozygous *TH* carriers are not known to be symptomatic, however a single case documents a heterozygous pathogenic mutation in an individual with levodopa-responsive dystonia^18^. Methods are provided in Supplementary Material: Genetics. Sequencing results in graphical form are provided in Supplemental Figure 1.

In idiopathic PD and in this PD patient with TH mutation, changes in DAT+ PBMCs were coupled to worsening motor function prior to carbidopa:levodopa therapy, and with improvement of UPDRS-III scores following carbidopa/levodopa treatment. Unlike idiopathic PD cases which we previously described^17^, changes in TH+ PBMCs and UPDRS-III scores were uncoupled. Consistent with previous reports, we found that in idiopathic PD carbidopa/levodopa treatment reduced the levels of these markers (Figure 2A-B). Therefore, in idiopathic PD, there was a direct correlation between changes in TH+ PBMCs, DAT+ PBMCs, carbidopa/levodopa and UPDRS-III scores. The immunophenotyping of a PD patient carrying a pathogenic TH mutation presented an opportunity to assess the association of the DAT and TH markers in this patient, and to compare them to an idiopathic PD cohort (n= 130) and to an age/sex matched healthy control cohort (n=80). As described above, the patient presented classic PD symptoms, and we hypothesized increased DAT+ and TH+ PBMCs at the time of PD diagnosis (drug naïve) would be higher than age/sex matched healthy subjects and would be coupled to improvement of UPDRS-III scores following carbidopa/levodopa treatment.

**Figure 2.**
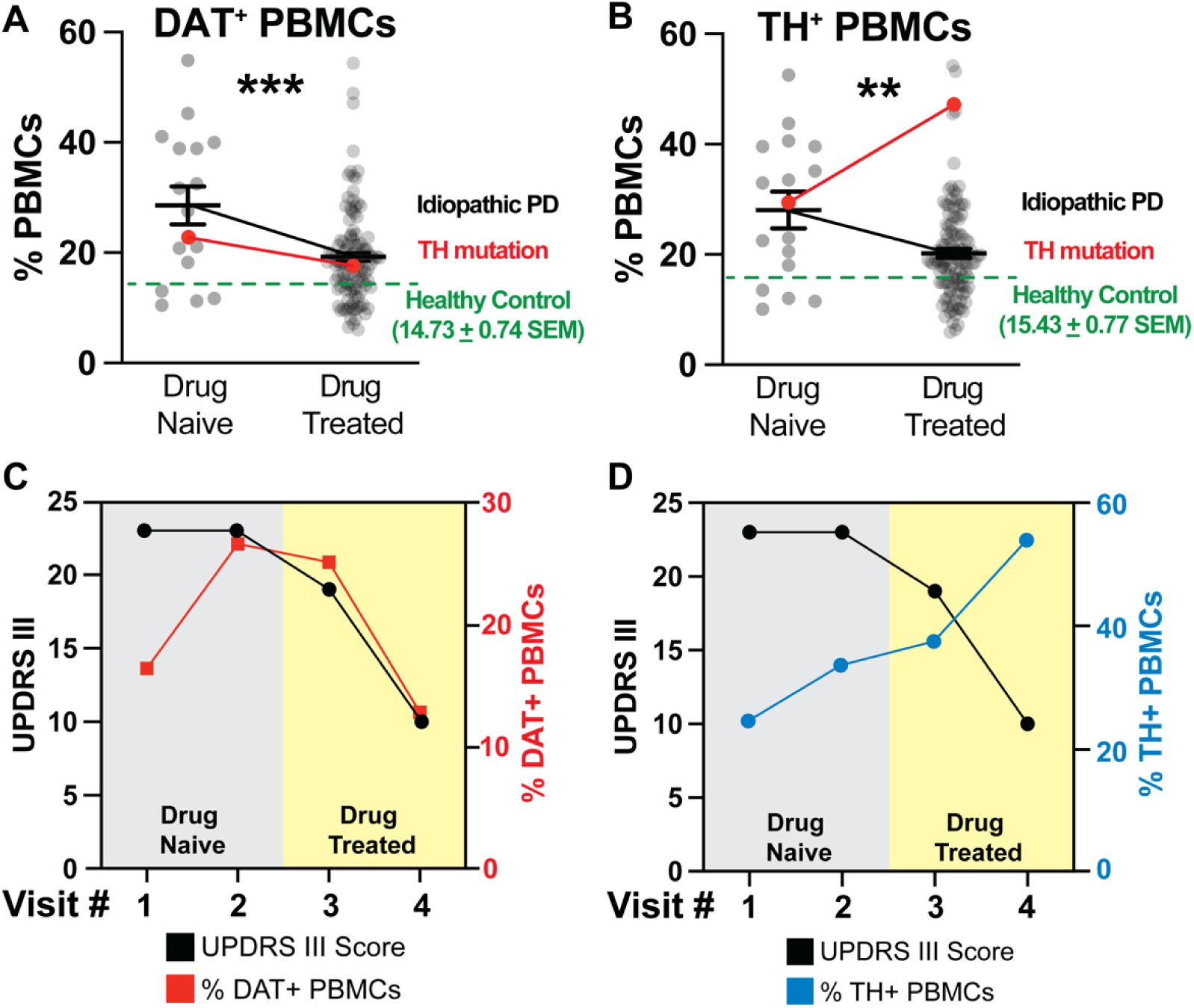
DAT+ and TH+ PBMCs are increased in idiopathic PD (idiopathic PD) patients at the time of diagnosis and decreased following improved UPDRS-III scores as a consequence of L-DOPA/Carbidopa treatment. By contrast, unlike the DAT+ PBMCs, in a patient carrying a TH mutation, the TH+ PBMCs remains elevated even after successful pharmacotherapy, representing an uncoupled response to L-DOPA/Carbidopa treatment. Single cells, free from debris, were analyzed via flow cytometry for expression of DAT and TH (Supplemental Figure 2). A-B) DAT+ and TH+ PBMCs (grey dots) are increased in drug naïve idiopathic PD patients (n=16) relative to healthy controls (n= 80, green dashed line). These markers are reduced significantly in drug treated idiopathic PD patients (n=114), though remaining significantly higher than age/sex matched healthy control levels (***p<0.001; t,df(3.763, 128), **p<0.01; t,df(3.138, 128), unpaired Student’s t-test with Tukey’s correction, alpha=0.05). In the PD patient carrying a single pathogenic TH mutation (red line), A) the pattern of DAT+ PBMCs change follows the pattern we identified in idiopathic PD patients, i.e., increased at the time of PD diagnosis (drug naïve condition) and then decreasing following successful L-DOPA/Carbidopa treatment. However, B) TH+ PBMCs of the PD patient carrying pathogenic TH mutation are uncoupled from both DAT+ PBMCs and the predicted reduction following L-DOPA/Carbidopa treatment, for instance, they are elevated in the drug naïve condition and continue to increase despite improved UPDRS-III, as a results of L-DOPA/Carbidopa treatment. C) Similar to idiopathic PD patients, the PD patient carrying pathogenic TH mutation, DAT+ PBMCs (red line) are increased at the first visit (drug naïve) and continue to increase as a function of worsening motor function (UPDRS-III, black line). Once L-DOPA/Carbidopa treatment is initiated, both DAT+ PBMCs and UPDRS-III scores decrease, indicating positive response to pharmacotherapy. D) In the PD patient carrying pathogenic TH mutation, the TH+ PBMCs (blue line) are increased following worsening motor symptoms (UPDRS-III, black line), but TH+ PBMCs continue to rise despite positive response to L-DOPA/Carbidopa (indicated by decreasing UPDRS-III motor scores), suggesting in this patient, only changes in the DAT+ PBMCs are coupled to both worsening motor function and positive L-DOPA/Carbidopa treatment response.

Consistent with our prior findings^17^, at the time of diagnosis (drug naïve), idiopathic PD patients (n=130) exhibited increased DAT+ and TH+ PBMCs (Figure 2A-B, grey dots, black line) relative to age/sex matched healthy control levels (green dashed line, n=80). idiopathic PD patients show a significant decrease in DAT+ and TH+ PBMCs, and improved UPDRS-III scores following carbidopa/levodopa treatment (Figure 2A-B, grey dots, black line, Figure 2A-B, green). Similar to idiopathic PD patients, the patient with TH mutation revealed an increased DAT+ PBMCs (Figure 2A, red line). Consistent with our hypothesis, carbidopa/levodopa treatment provided symptom relief marked by reduced UPDRS III scores (Figure 2C, black line) and was associated with a reduced frequency of DAT+ PBMCs (Figure 2C, red line). Thus, the dynamic changes in DAT+ PBMCs followed a similar pattern in this subject as in idiopathic PD cohort. Consistent with our hypothesis, in the patient with TH mutation, TH+ PBMCs were increased at the time of diagnosis (drug Naïve), but contrary to our hypothesis they remained high and continued to rise despite carbidopa/levodopa and the improved UPDRS-III scores (Figure 2B, red line, blue line, black line respectively). This finding was in contrast to that observed in idiopathic PD, where there was a direct correlation between decreased TH+ PBMC frequencies and improved UPDRS-III scores following carbidopa/levodopa treatment. The detailed methods are provided in the Supplementary Information: PBMC isolation and flow cytometry.

Unlike what is observed in a DaTscan, changes in DAT expressing peripheral immunophenotype reflect PD progression. In the patient with a TH mutation, the asymmetric reduction in DaTscan (Figure 1) at the time of PD diagnosis was reflected by an increase in DAT+ PBMCs (Figure 2) and UPDRS-III scores. However, the longitudinal DaTscan signal, measured 11 months later, was unchanged with no qualitatively differences observed between scans, despite the worsening of UPDRS-III scores, and the increase in DAT+ PBMC thus suggesting the peripheral immunophenotype may provide a more sensitive means to monitor disease progression.

## Concluding Remarks

The clinical manifestations, symptoms and progression of PD are continuous regardless of whether its origin is idiopathic or genetic^6,19,20^. The use of immune system-based approaches for diagnosing and monitoring PD is a relatively new and evolving field. There has been a considerable amount of large-scale and population-based research in this area, yet few longitudinal studies have systematically examined immunophenotyping in longitudinal PD cohorts^21–23^. Here we present a case of a PD patient with a heterozygous TH mutation. Longitudinal within subject sampling suggests that DAT+ PBMCs could serve as an indicator of disease progression, treatment response and as a potential diagnostic tool. We show that a TH gene variant can modulate TH-expression in peripheral immune cells and we posit that this may be causal or a consequence of PD in this single patient. As studies of other PD-associated gene variants have suggested^24–27^, genotype may affect the immunological profile in PD and should be taken into consideration when evaluating immunological tools for PD assessment.

Consistent with our previous study^17^ we documented that as UPDRS-III scores worsened, there was an elevation in DAT+ immune cells that was reduced after treatment with carbidopa/levodopa and there was subsequent improvement in UPDRS-III scores. The dynamic nature of DAT+ peripheral immune cells in the current study which was also conducted in idiopathic PD patients corroborate previous cross-sectional findings in idiopathic patients on this marker^17^. Intriguingly, in this PD patient harboring a heterozygous TH mutation, unlike DAT+ immune cells, the increase in TH+ immune cells persisted even following the positive response to carbidopa/levodopa. The current study marks the first instance of longitudinal immunophenotyping of a PD patient with TH mutation, highlighting DAT+ and TH+ PBMCs as potential indicators of PD, its progression, and the evolution of changes with treatment. Notably, the discernable disease progression and increased PD symptom severity, which was evident clinically, along with the changes of the DAT+ immunophenotype, were not reflected in two separate qualitative DaTscan’s collected 11 months apart. Prior studies have suggested an estimated annual decline in striatal DAT binding of -13% at the first year ^28^.

The limitations of the present study include the absence of longitudinal immunophenotyping and longitudinal DaTscan data for the idiopathic PD patient cohort allowing disease progression identification and treatment response across various patient subgroups. Another limitation is the absence of quantitative DaTscan data -due to technical limitation with the type of scan performed-on the patient with TH mutation. While the data presetned in this study suggest the identified PBMC immunophenotype can be used for disease detection, tracking progression of and response to pharmacotherapy in idiopathic PD patients, the inclusion of other genetic PD such as LRRK2 or GBA mutations may identify additional immunophenotypic differences. In conclusion, our findings underscore the notion that peripheral immunophenotype with regards to PBMCs expressing DAT offers a sensitive and robust tool for tracking disease onset, progression and therapeutic response in idiopathic PD and in a single case of a PD patient with TH mutation.

## Data Availability

Data are available upon request.

## Acknowledgements and funding

The Evelyn F. and William L. McKnight Brain Institute’s Gator NeuroScholar’s program (to AG), and the Karen Toffler Charitable Trust (to AG). In addition, these studies were accomplished using instrumentation and technical assistance from the University of Florida Center for Immunology and Transplantation.

## Supplementary Web Only Material

## Materials and Methods

### Human Subjects

Blood samples from age-matched healthy subjects were obtained from two sources: the Movement Disorder Clinic at the University of Florida *via* an approved IRB protocol with written informed consent (IRB201701195), or the Lifesouth Community Blood Center, Gainesville, FL where deidentified samples exempt from informed consent (IRB201700339) were purchased. According to Lifesouth regulations, healthy donors were individuals aged 40-80 years old of any gender, who were not known to have any blood borne pathogens (both self-reported and independently verified), and were never diagnosed with a blood disease, such as leukemia or bleeding disorders. In addition, none of the donors were using blood thinners or antibiotics, or were exhibiting signs/symptoms of infectious disease, or had a positive test for viral infection in the previous 21 days.

### Inclusion/exclusion criteria for human subjects

#### Parkinson’s disease patients

Potential study participants were evaluated by a board-certified neurologist specializing in movement disorders. Patients were eligible to participate if 1) they had a confirmed PD diagnosis, 2) there was absence of comorbid movement disorder (i.e. essential tremor), 3) there was absence of any psychiatric diagnoses, 4) they were not prescribed psychotropic medications (i.e. neuroleptics), 5) they had no current or recent diagnosis of cancer (within 18 months) and were not on current or recent (within 18 months) treatment for the same, 6) had not been diagnosed with viral, bacterial or other infections within the preceding 21 days and were currently not being treated for the same.

#### Healthy control subjects

While not evaluated explicitly by a movement disorder specialist, healthy control subjects were present at the time of blood draw for PD patients and most frequently included the patient’s spouse, allowing for control of environmental factors that may influence immune factors being studied. Participants were eligible to participate if 1) they report no current or past diagnosis of motor disorder (PD, ET, dystonia), 2) they were currently not taking medication for the same (self-reported), and 3) were not exhibiting overt symptoms of movement disorder.

#### Demographic information

Age, disease duration, sex distribution and motor scales for groups used for analysis are given in Supplemental Table 1.

### PBMC isolation and Flow cytometry

Materials and equipment are listed in Tables 1, 2 and 3. As previously published^29–32^, whole blood was collected in K2EDTA vacutainer blood collection tubes (BD, 366643) and kept at room temperature for up to 2 hours prior to PBMC isolation. Briefly, blood from healthy volunteers and PD patients was overlaid in Leucosep tubes for PBMC isolation, centrifuged for 20 minutes at 400g with brakes turned off and acceleration set to minimum. PBMCs were collected from the interphase of Ficoll and PBS, transferred to a fresh 15mL conical tube, resuspended in 8mL sterile PBS and centrifuged for 10 minutes at 100 x *g*, and repeated twice more. Cells were counted with a hemacytometer using trypan blue exclusion of dead cells, and density-adjusted with PBS for flow cytometry staining.

**Table 1:**
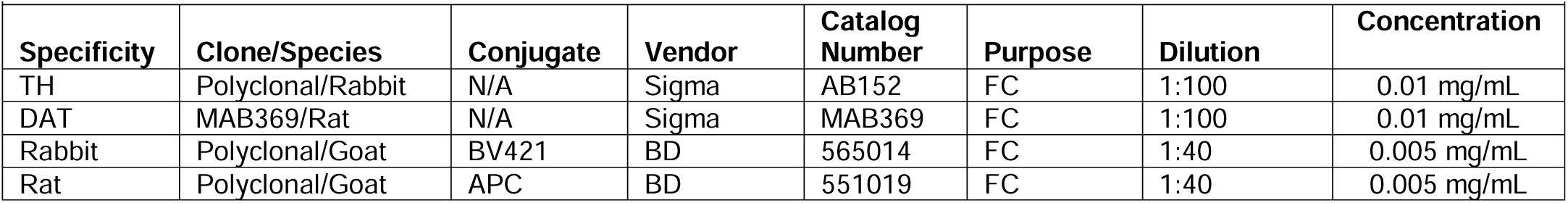
Antibodies.

| Specificity | Clone/Species | Conjugate | Vendor | Catalog Number | Purpose | Dilution | Concentration |
| --- | --- | --- | --- | --- | --- | --- | --- |
| TH | Polyclonal/Rabbit | N/A | Sigma | AB152 | FC | 1:100 | 0.01 mg/mL |
| DAT | MAB369/Rat | N/A | Sigma | MAB369 | FC | 1:100 | 0.01 mg/mL |
| Rabbit | Polyclonal/Goat | BV421 | BD | 565014 | FC | 1:40 | 0.005 mg/mL |
| Rat | Polyclonal/Goat | APC | BD | 551019 | FC | 1:40 | 0.005 mg/mL |

**Table 2:** Reagents and Materials.

| Reagent | Supplier | Catalog Number | Purpose | Concentration |
| --- | --- | --- | --- | --- |
| Ficoll-Paque Plus | GE | 45-001-750 | PBMC isolation | N/A |
| PBS | In house | N/A | PBMC isolation, FC | 1x |
| K2EDTA Vacutainer | BD | 366643 | Blood collection | N/A |
| Butterfly blood collection device | BD | 367342 | Blood collection | N/A |
| Leucosep Tubes | Grenier BioOne | 227290P | PBMC isolation | N/A |
| Trypan Blue | MP Biomedicals | 1691049 | Cell counting | Stock |
| Fix/Perm Kit | eBioscience | 88-8824-00 | FC | Stock |

**Table 3:** Equipment.

| Equipment | Supplier | Part Number | Purpose |
| --- | --- | --- | --- |
| Centrifuge | Sorvall | ST8 | PBMC isolation |
| Cytometer | BD | Canto II | FC |
| Spectral Analyzer | Cytek | Aurora | FC |
| Microcentrifuge | Fisher | 59A | FC |

Patient and healthy control subject PBMCs were stained for flow cytometry analysis in 100uL staining volume containing 1 million cells per condition. Staining for intracellular epitopes of DAT and TH was performed at room temperature in permeabilization buffer (eBioscience, 88-842-00), followed by species-specific secondary antibodies. We note that the flow cytometry method used to detect DAT and TH expressing cells does not allow for assessment of protein levels of these markers. Samples were resuspended in 500uL PBS after the final wash. Data were collected within 2 hours on BD Canto II or Cytek Aurora Spectral Cytometer. Each experiment included single color compensation, followed by automatic compensation calculation. Compensation matrices were not altered thereafter. Data were analyzed using Flowjo Software (BD Biosciences). Gating strategy is shown in Supplemental Figure 2.

#### Genetics

Samples and data is processed and stored according to HIPAA compliance requirements following CAP guidelines^33^ and CLIA standards for quality and competence^34^ at UF Health Medical Laboratories Whole genome sequencing (WGS) is benchmarked using the National Institute of Standards and Technology ‘Genome in a Bottle’ Consortium standards (HG002 son, HG003 & HG004 parental genomes^35^).WGS precision metrics have been explored from 5-30× depth for the entire genome and results are concordant with the NIST/PrecisionFDA data “truth sets”^36–38^ with > 95.2% analytical sensitivity and > 97.3% precision. DNA is extracted from a buccal swab using the Qiagen QIAmp DNA Mini Kit. DNA is quantified by fluorescence on an Invitrogen Qubit Fluorometer. Individually-indexed genomic libraries are prepared using dual unique indexes from ∼200ng DNA/individual (New England Biolabs NEBNext^®^ Ultra^TM^ II DNA Library Prep Kit for Illumina^®^). Genome library quality and quantity are confirmed by automated electrophoresis on an Agilent 2100 bioanalyzer and by qPCR. Individual libraries are normalized and pooled in equimolar ratios for 2× 150 bp paired-end sequencing at 35× depth on an Illumina NovaSeq 6000. To expedite and enable innovation, speed, data and code sharing, and to maintain security and PHI/HIPAA compliance, versioned bioinformatic pipelines for clinical genome variant calling have been developed in AWS in a containerized compute environment. Bioinformatic analyses include index deconvolution of pooled samples, read trimming, alignment, QC analyses, variant calling and annotation, with gene set panel filtering followed by variant prioritization by expert review^39–41^ In brief, .fastq reads are aligned to the human genome reference (GRCh37/hg19) and variants are called and annotated using ‘versioned’ open source softwares including: TrimmomaticPE 0.39^42^, FastQC 0.11.9, MultiQC 1.9^43^, BWA MEM 0.7.17-r1188^44^, samtools 1.10^45^, Picard 2.23.8^46^, Strelka2 2.9.10^47^, bcftools 1.10.2^45^, snpEff 5.0c^48^, ExpansionHunter 4.0.2^49^, Manta^50^ and cn.mops 1.8.0^51^. Computation is optimized by Nextflow^52^. Quality control reports are generated and examined for all individual samples and batched runs, including general statistics on WGS coverage per sample, on mapping quality and on the proportion of reads surviving that process. These data quantify sequence read counts (unique, duplicate and overrepresented %), quality (Phred scores across reads, per sequence quality scores, length distributions, GC content and ‘N’ scores). Aligned files (.bam, .bai) and annotated variant files include quality scores, read orientations and depths. Per sample variability is documented as a composite variant call file (VCF) that includes all intergenic regions, intronic variants, downstream and upstream gene variants, non-coding, missense, nonsense and silent/synonymous variants, frameshift, stop gain, splice, disruptive in-frame deletions and duplications, start loss, stop loss and gene fusions. Our annotation approach is exact, comprehensive^53,54^ and includes CADD^55^ and Revel scores^56^, gnomAD frequencies^57^, ClinVar^58^ and OMIM entries^59^. All ∼22,000 genes that make up the human genome are sequenced. Reporting is restricted to exonic nonsynonymous and splicing (± 20 bp) substitutions. Only variants with > 10× coverage are reported. The clinical significance of the filtered variants is assessed according to ACMG recommendations^40^. This sequence reported here is performed as research using de-identified swabs/DNA samples provided by collaborating investigators. However, this variant had been previously identified in this patient in a commercial CAP-accredited clinical lab.

#### DaTScan

DaTscan imaging was performed as published in Catafau 2004^60^. SPECT imaging was obtained 3 to 6 hours following intravenous injection of ^123^I-Ioflupane (111-185 MBq; DaTscan GE Healthcare, Amersham, UK). A dose of 4.36 mCi of I-123 ioflupane were utilized. Images were acquired using a gamma camera fitted with high-resolution collimators and set to a photopeak of 159 keV with a ± 10% energy window. Subject was supine with the head on an off-the-table headrest, a flexible head restraint such as a strip of tape across the chin or forehead may be used to help avoid movement and set a circular orbit for the detector heads with the radius as small as possible. Interpretation of results indicated asymmetric uptake and activity [e.g. activity in the region of the putamen of one hemisphere is absent or greatly reduced with respect to the other] and reported as abnormal results. There was reduced activity in both right and left putamen alone or also in the caudate nuclei.

### Statistics

Unpaired Student’s t-test (two-tailed) was used when comparing two groups with Tukey’s correction for multiple comparisons. P<0.05 was considered statistically significant. using unpaired Student’s t-test (two-tailed) All statistical analyses were performed in GraphPad Prism 10.

---

**Supplemental Figure 1.**
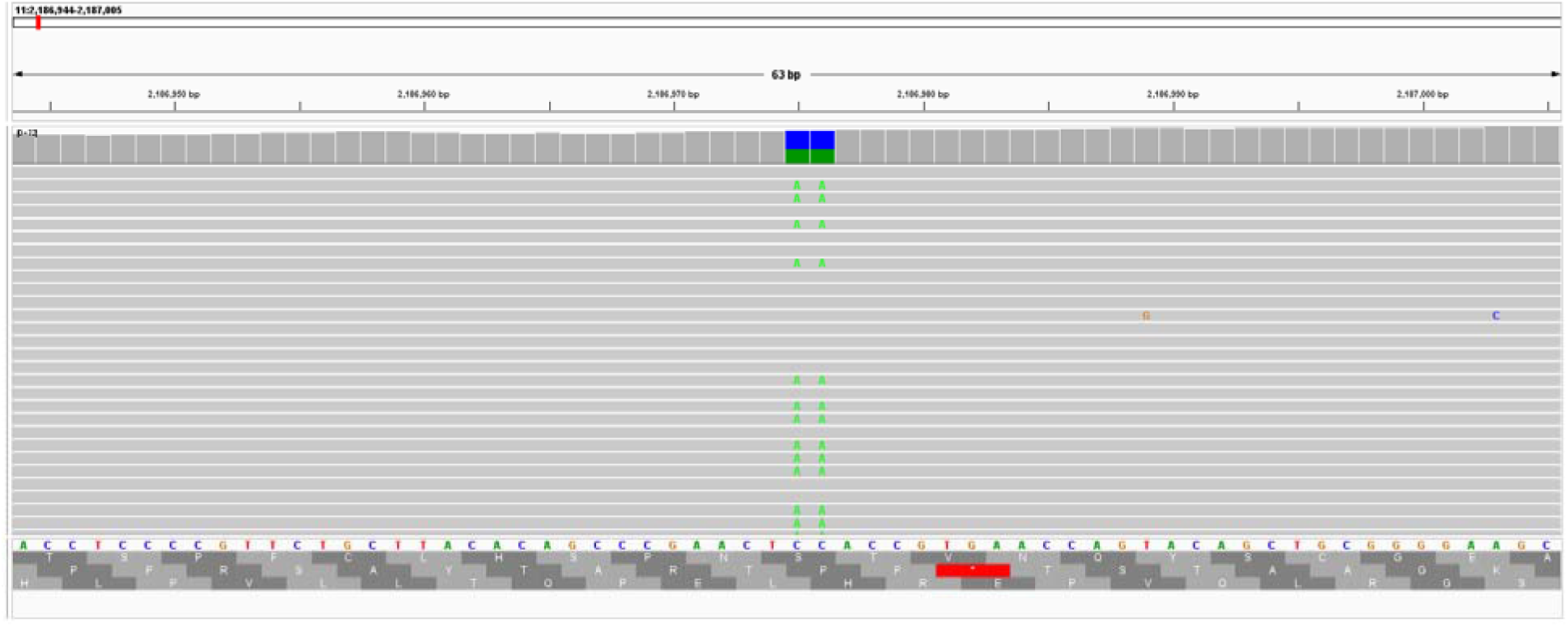
Sequencing results. Integrated genome view of chromosome 11: 2,186,944-2,187,005 interval spanning the TH locus, exon 12, and the position of the g.2,186,975-6 CC>AA (c.1215_1216delCCinsTT; p. Glu406*) pathogenic variant. Reads and their orientation are illustrated in gray, the consensus nucleotide sequence is shown beneath, along with possible open reading frames and the translated protein normally encoded (blue bar). ‘CTC’ encoding “E”, the single letter amino acid for glutamine (Glu) is mutated to ‘ATC’ encoding the amber stop codon.

**Supplemental Figure 2.**
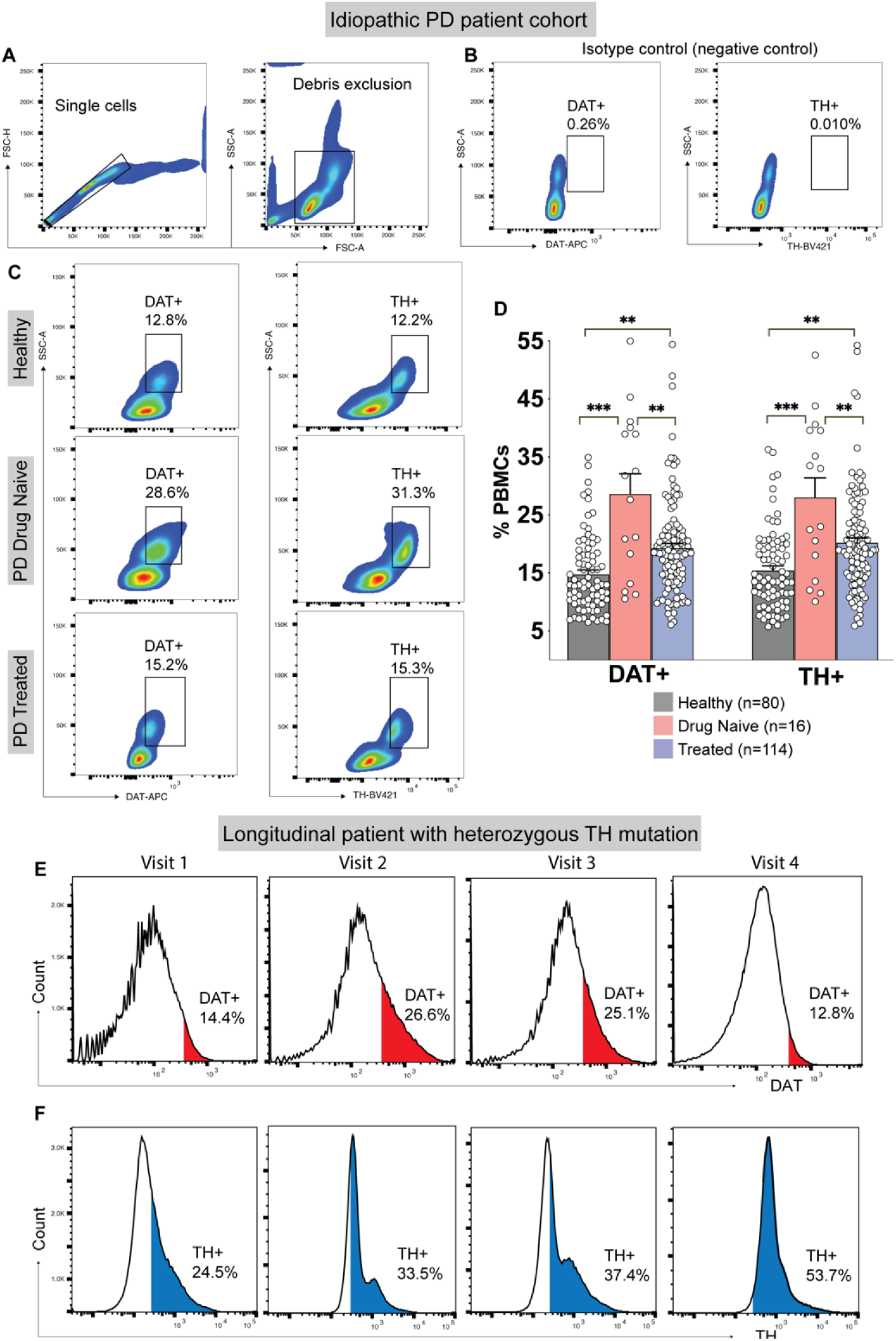
Gating strategy and representative DAT+ and TH+ PBMCs in a cohort of idiopathic PD patients, healthy control subjects, and a PD patient carrying a TH mutation. Isolated PBMCs were stained and analyzed via flow cytometry after A) gating single cells and excluding debris. B) Non-immune isotype control was used to assess specificity of DAT and TH signals. C-D) Relative to healthy control subjects, drug naïve idiopathic PD patients show a significantly increased percentage of DAT+ and TH+ PBMCs, which are significantly reduced in treated patients, but remain higher than healthy control levels (**p<0.01, ***p<0.001; One-way ANOVA, with Tukey’s correction for multiple comparisons; alpha=0.05; f(2,3)=305.7). E) In a patient carrying a heterozygous TH mutation, DAT+ PBMCs follow the trend established in idiopathic PD, where DAT+ PBMCs are increased prior to treatment, and subsequently decrease towards healthy control levels (Figure 3) following treatment. In contrast to idiopathic PD patients, F) the subject carrying a TH mutation shows TH+ PBMCs continuing to increase despite treatment for PD, indicating TH+ PBMCs are uncoupled to treatment response.

**Supplemental Table 4:** Patient demographics.

| # Female | # Male |  | Disease Duration (Years) | Age (Years) | HY Score* | UPDRS-III (Off)* | UPDRS-III (On)* | LED* |
| --- | --- | --- | --- | --- | --- | --- | --- | --- |
| 47 | 83 | Average | 10.66 | 67.72 | 2.51 | 31.03 | 24.46 | 1111.81 |
|  |  | Range | 0-27 | 41-81 | 1.5-5 | 13-67 | 2-51 | 0-25000 |
|  |  | Standard Deviation | 7.17 | 7.99 | 0.75 | 10.95 | 9.83 | 2252.61 |

\*HY = Hoehn and Yahr score; UPDRS-III = Unified Parkinson’s Disease Rating Scale part 3, motor subscale; On and Off refer to on drug or off drug status; LED = Levodopa Equivalence Dose

## References

1 Ou, Z. et al. Global Trends in the Incidence, Prevalence, and Years Lived With Disability of Parkinson’s Disease in 204 Countries/Territories From 1990 to 2019. Front Public Health 9, 776847, doi:10.3389/fpubh.2021.776847 (2021).

2 Deng, H., Wang, P. & Jankovic, J. The genetics of Parkinson disease. Ageing Res Rev 42, 72–85, doi:10.1016/j.arr.2017.12.007 (2018).

3 Blauwendraat, C., Nalls, M. A. & Singleton, A. B. The genetic architecture of Parkinson’s disease. Lancet Neurol 19, 170–178, doi:10.1016/s1474-4422(19)30287-x (2020).

4 Jia, F., Fellner, A. & Kumar, K. R. Monogenic Parkinson’s Disease: Genotype, Phenotype, Pathophysiology, and Genetic Testing. Genes (Basel) 13, doi:10.3390/genes13030471 (2022).

5 Guadagnolo, D., Piane, M., Torrisi, M. R., Pizzuti, A. & Petrucci, S. Genotype-Phenotype Correlations in Monogenic Parkinson Disease: A Review on Clinical and Molecular Findings. Frontiers in Neurology 12, doi:10.3389/fneur.2021.648588 (2021).

6 Correia Guedes, L., Mestre, T., Outeiro, T. F. & Ferreira, J. J. Are genetic and idiopathic forms of Parkinson’s disease the same disease? J Neurochem 152, 515–522, doi:10.1111/jnc.14902 (2020).

7 Tolosa, E., Garrido, A., Scholz, S. W. & Poewe, W. Challenges in the diagnosis of Parkinson’s disease. Lancet Neurol 20, 385–397, doi:10.1016/s1474-4422(21)00030-2 (2021).

8 Heim, B., Krismer, F., De Marzi, R. & Seppi, K. Magnetic resonance imaging for the diagnosis of Parkinson’s disease. J Neural Transm (Vienna) 124, 915–964, doi:10.1007/s00702-017-1717-8 (2017).

9 Doppler, K. Detection of Dermal Alpha-Synuclein Deposits as a Biomarker for Parkinson’s Disease. J Parkinsons Dis 11, 937–947, doi:10.3233/jpd-202489 (2021).

10 Gibbons, C. et al. Cutaneous α-Synuclein Signatures in Patients With Multiple System Atrophy and Parkinson Disease. Neurology 100, e1529–e1539, doi:10.1212/wnl.0000000000206772 (2023).

11 Pitton Rissardo, J. & Fornari Caprara, A. L. Cardiac 123I-Metaiodobenzylguanidine (MIBG) Scintigraphy in Parkinson’s Disease: A Comprehensive Review. Brain Sciences 13, 1471 (2023).

12 Skowronek, C., Zange, L. & Lipp, A. Cardiac 123I-MIBG Scintigraphy in Neurodegenerative Parkinson Syndromes: Performance and Pitfalls in Clinical Practice. Front Neurol 10, 152, doi:10.3389/fneur.2019.00152 (2019).

13 Catalan, M. et al. 123I-Metaiodobenzylguanidine Myocardial Scintigraphy in Discriminating Degenerative Parkinsonisms. Movement Disorders Clinical Practice 8, 717–724, 10.1002/mdc3.13227 (2021).

14 Lee, J. Y. et al. Multimodal brain and retinal imaging of dopaminergic degeneration in Parkinson disease. Nat Rev Neurol 18, 203–220, doi:10.1038/s41582-022-00618-9 (2022).

15 Qi, R. et al. A blood-based marker of mitochondrial DNA damage in Parkinson’s disease. Science Translational Medicine 15, eabo1557, doi:doi:10.1126/scitranslmed.abo1557 (2023).

16 Di Maio, R. et al. LRRK2 activation in idiopathic Parkinson’s disease. Science Translational Medicine 10, eaar5429, doi:doi:10.1126/scitranslmed.aar5429 (2018).

17 Gopinath, A. et al. DAT and TH expression marks human Parkinson’s disease in peripheral immune cells. NPJ Parkinsons Dis 8, 72, doi:10.1038/s41531-022-00333-8 (2022).

18 Bally, J. F. et al. Mild dopa-responsive dystonia in heterozygous tyrosine hydroxylase mutation carrier: Evidence of symptomatic enzyme deficiency? Parkinsonism Relat Disord 71, 44–45, doi:10.1016/j.parkreldis.2020.01.017 (2020).

19 Jankovic, J. & Tan, E. K. Parkinson’s disease: etiopathogenesis and treatment. J Neurol Neurosurg Psychiatry 91, 795–808, doi:10.1136/jnnp-2019-322338 (2020).

20 Schiesling, C., Kieper, N., Seidel, K. & Krüger, R. Review: Familial Parkinson’s disease--genetics, clinical phenotype and neuropathology in relation to the common sporadic form of the disease. Neuropathol Appl Neurobiol 34, 255–271, doi:10.1111/j.1365-2990.2008.00952.x (2008).

21 Espinosa-Cárdenas, R. et al. Immunomodulatory effect and clinical outcome in Parkinson’s disease patients on levodopa-pramipexole combo therapy: A two-year prospective study. J Neuroimmunol 347, 577328, doi:10.1016/j.jneuroim.2020.577328 (2020).

22 Lindestam Arlehamn, C. S., et al. α-Synuclein-specific T cell reactivity is associated with preclinical and early Parkinson’s disease. Nat Commun 11, 1875, doi:10.1038/s41467-020-15626-w (2020).

23 Williams-Gray, C. H. et al. Serum immune markers and disease progression in an incident Parkinson’s disease cohort (ICICLE-PD). Mov Disord 31, 995–1003, doi:10.1002/mds.26563 (2016).

24 Al-Azzawi, Z. A. M., Arfaie, S. & Gan-Or, Z. GBA1 and The Immune System: A Potential Role in Parkinson’s Disease? J Parkinsons Dis 12, S53–s64, doi:10.3233/jpd-223423 (2022).

25 Kung, P. J., Elsayed, I., Reyes-Pérez, P. & Bandres-Ciga, S. Immunogenetic Determinants of Parkinson’s Disease Etiology. J Parkinsons Dis 12, S13–s27, doi:10.3233/jpd-223176 (2022).

26 Tansey, M. G. et al. Inflammation and immune dysfunction in Parkinson disease. Nat Rev Immunol 22, 657–673, doi:10.1038/s41577-022-00684-6 (2022).

27 Gate, D. New Perspectives on Immune Involvement in Parkinson’s Disease Pathogenesis. J Parkinsons Dis 12, S5–s11, doi:10.3233/jpd-223240 (2022).

28 Ikeda, K., Ebina, J., Kawabe, K. & Iwasaki, Y. Dopamine Transporter Imaging in Parkinson Disease: Progressive Changes and Therapeutic Modification after Anti-parkinsonian Medications. Intern Med 58, 1665–1672, doi:10.2169/internalmedicine.2489-18 (2019).

29 Mackie, P. M. et al. Functional characterization of the biogenic amine transporters on human macrophages. JCI Insight 7, doi:10.1172/jci.insight.151892 (2022).

30 Gopinath, A. et al. A novel approach to study markers of dopamine signaling in peripheral immune cells. J Immunol Methods 476, 112686, doi:10.1016/j.jim.2019.112686 (2020).

31 Gopinath, A. et al. TNFα increases tyrosine hydroxylase expression in human monocytes. NPJ Parkinsons Dis 7, 62, doi:10.1038/s41531-021-00201-x (2021).

32 Mackie, P. M. et al. Functional characterization of the biogenic amine transporter system on human macrophages. bioRxiv, 2021.2009.2008.459459, doi:10.1101/2021.09.08.459459 (2021).

33 Roy, S. et al. Standards and Guidelines for Validating Next-Generation Sequencing Bioinformatics Pipelines: A Joint Recommendation of the Association for Molecular Pathology and the College of American Pathologists. J Mol Diagn 20, 4–27, doi:10.1016/j.jmoldx.2017.11.003 (2018).

34 Schneider, F., Maurer, C. & Friedberg, R. C. International Organization for Standardization (ISO) 15189. Ann Lab Med 37, 365–370, doi:10.3343/alm.2017.37.5.365 (2017).

35 Genome in a bottle—a human DNA standard. Nature Biotechnology 33, 675–675, doi:10.1038/nbt0715-675a (2015).

36 Krusche, P. et al. Best practices for benchmarking germline small-variant calls in human genomes. Nat Biotechnol 37, 555–560, doi:10.1038/s41587-019-0054-x (2019).

37 Zook, J. M. et al. Extensive sequencing of seven human genomes to characterize benchmark reference materials. Sci Data 3, 160025, doi:10.1038/sdata.2016.25 (2016).

38 Zook, J. M. et al. A robust benchmark for detection of germline large deletions and insertions. Nat Biotechnol 38, 1347–1355, doi:10.1038/s41587-020-0538-8 (2020).

39 Kalia, S. S. et al. Recommendations for reporting of secondary findings in clinical exome and genome sequencing, 2016 update (ACMG SF v2.0): a policy statement of the American College of Medical Genetics and Genomics. Genet Med 19, 249–255, doi:10.1038/gim.2016.190 (2017).

40 Richards, S. et al. Standards and guidelines for the interpretation of sequence variants: a joint consensus recommendation of the American College of Medical Genetics and Genomics and the Association for Molecular Pathology. Genet Med 17, 405–424, doi:10.1038/gim.2015.30 (2015).

41 Riggs, E. R. et al. Technical standards for the interpretation and reporting of constitutional copy-number variants: a joint consensus recommendation of the American College of Medical Genetics and Genomics (ACMG) and the Clinical Genome Resource (ClinGen). Genet Med 22, 245–257, doi:10.1038/s41436-019-0686-8 (2020).

42 Bolger, A. M., Lohse, M. & Usadel, B. Trimmomatic: a flexible trimmer for Illumina sequence data. Bioinformatics 30, 2114–2120, doi:10.1093/bioinformatics/btu170 (2014).

43 Ewels, P., Magnusson, M., Lundin, S. & Käller, M. MultiQC: summarize analysis results for multiple tools and samples in a single report. Bioinformatics 32, 3047–3048, doi:10.1093/bioinformatics/btw354 (2016).

44 Li, H. Aligning sequence reads, clone sequences and assembly contigs with BWA-MEM. arXiv 1303.3997, 10.48550/arXiv.1303.3997 (2013).

45 Danecek, P. et al. Twelve years of SAMtools and BCFtools. Gigascience 10, doi:10.1093/gigascience/giab008 (2021).

46 Institute, B. Picard Toolkit. Github Repository (2019).

47 Kim, S. et al. Strelka2: fast and accurate calling of germline and somatic variants. Nat Methods 15, 591–594, doi:10.1038/s41592-018-0051-x (2018).

48 Cingolani, P. et al. A program for annotating and predicting the effects of single nucleotide polymorphisms, SnpEff: SNPs in the genome of Drosophila melanogaster strain w1118; iso-2; iso-3. Fly (Austin) 6, 80–92, doi:10.4161/fly.19695 (2012).

49 Dolzhenko, E. et al. ExpansionHunter: a sequence-graph-based tool to analyze variation in short tandem repeat regions. Bioinformatics 35, 4754–4756, doi:10.1093/bioinformatics/btz431 (2019).

50 Chen, X. et al. Manta: rapid detection of structural variants and indels for germline and cancer sequencing applications. Bioinformatics 32, 1220–1222, doi:10.1093/bioinformatics/btv710 (2015).

51 Povysil, G., et al. panelcn.MOPS: Copy-number detection in targeted NGS panel data for clinical diagnostics. Hum Mutat 38, 889–897, doi:10.1002/humu.23237 (2017).

52 Di Tommaso, P. et al. Nextflow enables reproducible computational workflows. Nature Biotechnology 35, 316–319, doi:10.1038/nbt.3820 (2017).

53 Ejigu, G. F. & Jung, J. Review on the Computational Genome Annotation of Sequences Obtained by Next-Generation Sequencing. Biology (Basel) 9, doi:10.3390/biology9090295 (2020).

54 Zerbino, D. R., Frankish, A. & Flicek, P. Progress, Challenges, and Surprises in Annotating the Human Genome. Annu Rev Genomics Hum Genet 21, 55–79, doi:10.1146/annurev-genom-121119-083418 (2020).

55 Kircher, M. et al. A general framework for estimating the relative pathogenicity of human genetic variants. Nat Genet 46, 310–315, doi:10.1038/ng.2892 (2014).

56 Ioannidis, N. M. et al. REVEL: An Ensemble Method for Predicting the Pathogenicity of Rare Missense Variants. Am J Hum Genet 99, 877–885, doi:10.1016/j.ajhg.2016.08.016 (2016).

57 Karczewski, K. J. et al. The mutational constraint spectrum quantified from variation in 141,456 humans. Nature 581, 434–443, doi:10.1038/s41586-020-2308-7 (2020).

58 Landrum, M. J. et al. ClinVar: improving access to variant interpretations and supporting evidence. Nucleic Acids Res 46, D1062–d1067, doi:10.1093/nar/gkx1153 (2018).

59 Amberger, J., Bocchini, C. A., Scott, A. F. & Hamosh, A. McKusick’s Online Mendelian Inheritance in Man (OMIM). Nucleic Acids Res 37, D793–796, doi:10.1093/nar/gkn665 (2009).

60 Catafau, A. M. & Tolosa, E. Impact of dopamine transporter SPECT using 123I-Ioflupane on diagnosis and management of patients with clinically uncertain Parkinsonian syndromes. Mov Disord 19, 1175–1182, doi:10.1002/mds.20112 (2004).

